# Speech cortical activation and connectivity in typically developing children and those with listening difficulties

**DOI:** 10.1101/2020.10.26.20218495

**Authors:** Hannah J. Stewart, Erin K. Cash, Lisa L. Hunter, Thomas Maloney, Jennifer Vannest, David R. Moore

## Abstract

Listening difficulties (LiD) in people who have normal audiometry are a widespread but poorly understood form of hearing impairment. Recent research suggests that childhood LiD are cognitive rather than auditory in origin. We examined decoding of sentences using a novel combination of behavioral testing and fMRI with 43 typically developing children and 42 age matched (6-13 years old) children with LiD, categorized by caregiver report (ECLiPS). Both groups had clinically normal hearing. For sentence listening tasks, we found no group differences in fMRI brain cortical activation by increasingly complex speech stimuli that progressed in emphasis from phonology to intelligibility to semantics. Using resting state fMRI, we examined the temporal connectivity of cortical auditory and related speech perception networks. We found significant group differences only in cortical connections engaged when processing more complex speech stimuli. The strength of the affected connections was related to the children’s performance on tests of dichotic listening, speech-in-noise, attention, memory and verbal vocabulary. Together, these results support the novel hypothesis that childhood LiD reflects difficulties in language rather than in auditory or phonological processing.

## INTRODUCTION

Listening is commonly defined as active attention to sound. In human communication, listening also involves memory, language and executive function (Rudner & Signoret, 2016; Schiller et al., 2022). Listening difficulties (LiD) are frequently reported by caregivers of children with clinically normal hearing (Petley et al., 2021; Roeser et al., 2007). However, the mechanisms underlying childhood LiD without hearing loss remain poorly understood. Several studies have shown that such children typically also have a variety of academic, speech, language, attention and other developmental learning problems (Ferguson et al., 2011; Moore et al., 2018; Sharma et al., 2009). Some of these children receive a clinical diagnosis of auditory processing disorder (APD; American Academy of Audiology, 2010; British Society of Audiology, 2018; Dawes & Bishop, 2009).

The heterogeneous nature of these children’s difficulties has led to considerable controversy surrounding APD (Iliadou et al., 2019; Iliadou et al., 2018; McFarland & Cacace, 2009; Moore, 2018; Neijenhuis et al., 2019; Rees, 1973). At the center of this controversy is the question of whether the children have primarily auditory sensory problems (“bottom-up”), cognitive and language problems (“top-down”), or some combination of the two (Dillon & Cameron, 2021; Moore et al., 2010). In this and other recent studies (Hunter et al., 2021; Petley et al., 2021) we sidestep this controversy by using the umbrella term LiD (Dillon & Cameron, 2021), and the quantitative metrics of a well-validated, reliable and standardized caregiver questionnaire (ECLiPS; Barry & Moore, 2021) to operationalize LiD.

Speech understanding requires the coalescence of bottom-up processing of acoustic features in the auditory pathway, with top-down processing of linguistic features in speech and language pathways. These top-down processing pathways are thought to include frontal, temporal and parietal cortices involved in semantic representation, memory and attention (Hickok & Poeppel, 2007; Pichora-Fuller et al., 2016; Rönnberg et al., 2019; B. G. Shinn-Cunningham, 2008). Impaired interactions between the auditory system and these higher-level cortical speech perception systems could hold the key to further understanding of how LiD presents in children. Poor performance on complex auditory tasks (e.g. speech listening in noise) has been more closely associated with cognitive function than with performance on simpler, non-speech based auditory tasks (Moore et al., 2010).

Speech morphs into language as it progresses through a hierarchy of processing competencies aeading to comprehension. First, auditory information is classified into linguistically meaningful units called phonemes. This stage bridges bottom-up and top-down processing (Brodbeck et al., 2018). Second, the listener links phonemes into words to judge the intelligibility of the auditory information by comparing the words to memory templates. Third, meaning (semantics) is attached to one or more intelligible words. Basic and speech-related acoustic cues and features enable each of these processing competencies. For example, pitch, loudness, temporal order are required to distinguish individual phonemes. Speech-related features (e.g. voicing, vowel quality, direction of formant transition and noise burst cues) aid in linking the phonemes into words and words into sentences.

Scott et al. (2000) designed a paradigm, subsequently modified by Halai et al. (2015), to investigate cortical activity produced by different levels of speech processing competencies in adults (Figure 1A). They used Clear speech and two types of speech-like stimuli: Rotated speech, that spectrally rotated the stimuli around 2 kHz (Blesser, 1972); and Rotated+Vocoded speech, that first rotated and then noise vocoded the stimuli into 6 frequency bands (Shannon et al., 1995). In each of the three stimulus types, the sequential order of phonemes was preserved, thus retaining the prosody (i.e. rhythm) from the Clear speech. In the Rotated speech, the non-spectral features used to identify phonemes remained, while the spectral features were transformed. For example, the formant transition and noise burst cues that distinguish consonants were shifted to new frequencies and so were processed by different regions of the cochlea. This removed the intelligibility of the stimuli while maintaining the prosodic and phonetic features. Finally, in the Rotated+Vocoded speech, the intonation and phonetic features were removed, along with intelligibility.

**Figure 1:**
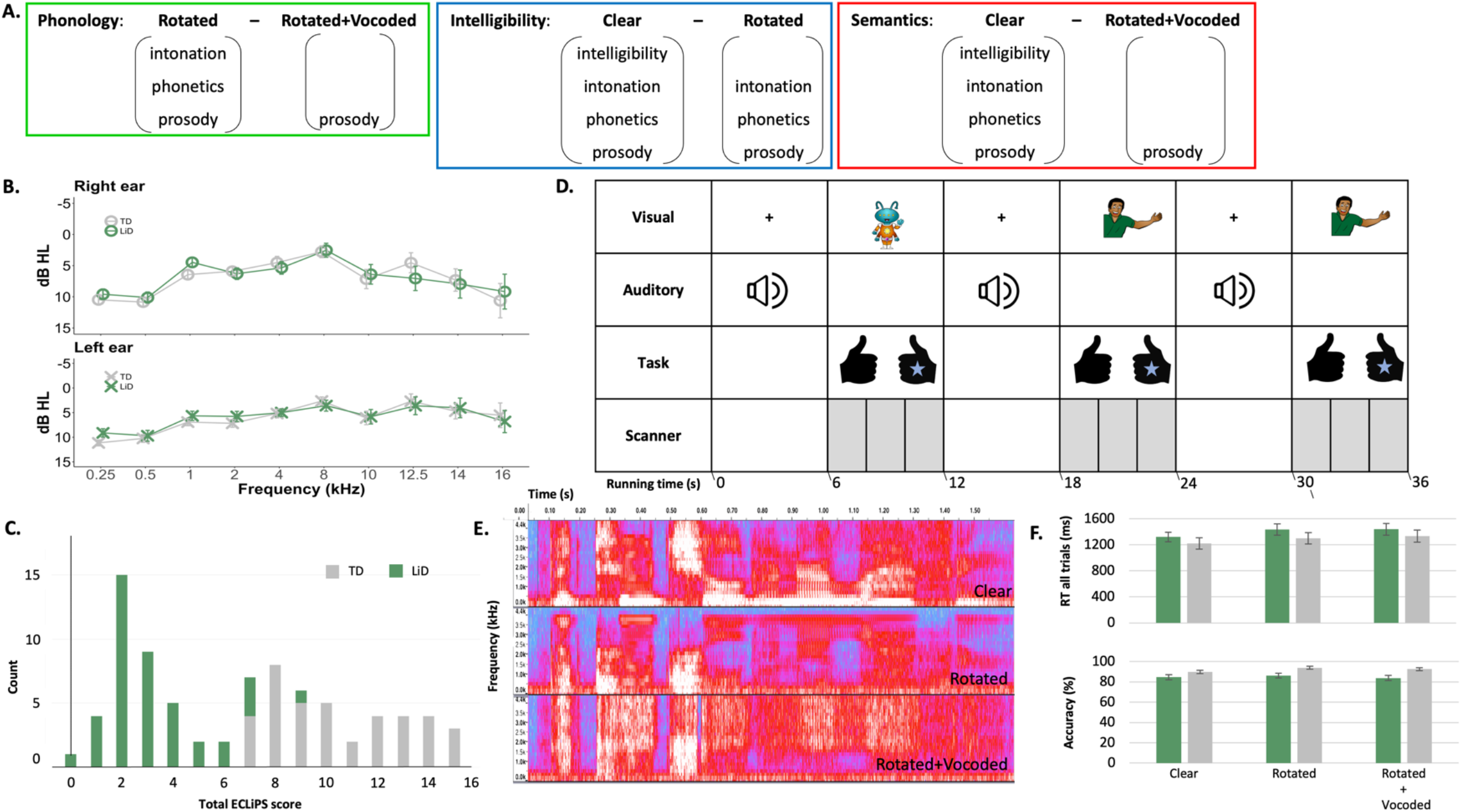
All children had normal tone hearing and were able to perform the speech perception task in the scanner. (A) Stimulus contrasts, illustrating how subtracting one stimulus type from another (Fig. 1E) isolates the specific speech processing competencies of Phonology, Intelligibility and Semantics. (B) Group mean hearing thresholds. (C) Total ECLiPS score. (D) fMRI paradigm. Children were asked if the picture (alien, man) matched who spoke the sentence (stimulus type, Fig. 1E). If it was a match they pressed the right button (with their stickered hand), if it was not a match they pressed the left button. Data acquisition (grey shading) was turned off/on for the presentation of the auditory stimuli (HUSH/sparse scanning). (E) Stimulus types. Spectrograms of ‘The two children are laughing’. Time is represented on the x-axis (0.0 –1.60 s) and frequency on the y-axis (0.0–4.4 kHz). The shading of the trace in each time/frequency region is controlled by the amount of energy in the signal at that particular frequency and time (red = more energy, blue = less energy). Clear speech (“man”) is intelligible with intonation. Rotated speech (“alien”) is not intelligible, though phonetic features and original intonation are preserved. Rotated + Vocoded speech (“alien”) is completely unintelligible, but preserves the character of the envelope and some spectral detail. (F) Group mean response times and accuracy for each stimulus type. Error bars represent the standard error of the mean. Groups as in Fig. 1C.

To explore which cortical areas were recruited for each of the three speech processing competencies, Scott et al. (2000) created analysis contrasts by subtracting sentence types from one another (Figure 1A). First, phonology was assessed by contrasting Rotated with Rotated+Vocoded sentences. Second, the judgement of intelligibility was assessed by contrasting Clear sentences with Rotated sentences. Finally, the ability to attach meaning to the intelligible sentence was assessed by contrasting Clear sentences with Rotated+Vocoded sentences where the acoustic complexity was matched but all meaning was removed.

In this study we adopted the methods developed by Scott et al. (2000) to identify cortical areas used in different levels of speech processing. Consistent with a top-down, linguistic model, and previous research in adults (Halai et al., 2015; Scott et al., 2000), we hypothesised that there would be no group difference in primary auditory cortex (i.e., Heschl’s gyrus) between the three contrasts of speech processing competencies. However, Intelligibility and Semantic contrasts would show less activation of the speech processing areas (e.g. superior temporal gyrus, Wernicke’s) in children with LiD than in TD children. It was expected that the spread of cortical activation for the three contrasts would increase from Phonology to Intelligibility to Semantics, and would include overlap in auditory and speech areas.

We next used the task-based cortical activation results to create regions-of-interest (ROIs) for functional connectivity analysis of a separate rs-fMRI acquired in the same scanning session. A rs-fMRI scan allows measurement of the temporal correlation of non-stimulus evoked fluctuations in the BOLD signal between anatomically separated brain regions (Biswal, 2012), capturing interactions between regions in functionally associated networks (Damoiseaux et al., 2006). We hypothesised that children with LiD would show diminished functional connectivity in cortical speech processing networks compared with TD children.

The aim of the study was to examine the cortical networks involved in different levels of speech processing in children with LiD relative to TD children. Our first research question asked if the two groups of children (LiD and TD) differ in cortical activation during speech listening, specifically in early auditory and speech processing cortical areas. Our second question investigated how the cortical areas activated during these speech processing competencies work together and how they relate to behavioural assessments of speech in noise identification, dichotic listening and cognition.

## RESULTS

We present here data from the first wave of a longitudinal study investigating the audiological and cognitive abilities of children (6-13 years old; Table 1) with LiD and their typically developing peers (6-13 years old; Table 1; Petley et al., 2021). All participants had clinically normal audiometry (≤ 20 dB HL, bilaterally; 0.25 – 8 kHz, Figure 1B), tympanometry (Hunter et al., 2021) and afferent auditory brainstem function (Hunter et al., 2022). Participants who scored at or below the bottom 10^th^ percentile on the total score of a caregiver checklist of everyday listening skills, the ECLiPS (Barry et al., 2015; Barry & Moore, 2021; Roebuck & Barry, 2018), were classified as having LiD (Figure 1C).

**Table 1:** Participant details for each scan type – fMRI speech listening task and resting state (RS). * High school graduate or less, some college or more

| Scan type | Group | N | Age M (SD) | Gender M, F | Maternal education* | Handedness L, both, R | History of tubes | Motion-related artifacts M (SD) |
| --- | --- | --- | --- | --- | --- | --- | --- | --- |
| fMRI | LiD | 43 | 10.02 (2.13) | 30, 13 | 6, 27 | 3, 5, 35 | 12 | 12.16 (4.63) |
|  | TD | 42 | 9.78 (1.93) | 25, 17 | 0, 42 | 2, 1, 39 | 15 | 11.67 (4.30) |
| RS | LiD | 42 | 10.06 (2.09) | 29, 13 | 8, 34 | 3, 6, 33 | 12 | 28.64 (32.17) |
|  | TD | 39 | 9.75 (1.93) | 23, 16 | 0, 39 | 2, 1, 36 | 15 | 23.00 (29.20) |

### Task-based cortical activation

During task-based MRI acquisition intervals (Figure 1D), children listened to Clear (“man”) or distorted (“alien”; Rotated and Rotated+Vocoded) spoken sentence types (Figure 1E) that we asked them to match to a visual cartoon representation. All children responded quickly and accurately on the sentence recognition task (Figure 1F), suggesting that they maintained attention throughout the task. Groups did not differ on reaction time (F(1, 83) = .89, p = .35, ηp2 = .011), but TD children were more accurate than children with LiD (F(1, 83) = 8.77, p = .004, ηp2 = .096). Sentence type did not affect response accuracy (F(1.78, 148.03) = 2.11, p = .13, ηp2 = .025) but did influence reaction time (F(2, 166) = 11.50, p < .001, ηp2 = .12) with Clear sentences eliciting quicker responses than Rotated sentences (p < .001, d = -.41) and Rotated+Vocoded sentences (p < .001, d = -.46). There were no significant interactions.

The two groups did not differ significantly between contrasts (Phonology, Intelligibility, Semantics; Figure 1A) after correcting for multiple comparisons (Figure 2A-C, clustering threshold = 2.3 voxels, family wise error (FWE) = .95). Further analysis of contrasts was therefore averaged across all children (n = 85; Figure 2D-F). The Phonology contrast (Figure 2D) showed bilateral activation in the middle and superior temporal gyrus, including Heschl’s gyrus, and temporal pole. Activation was also found in the left hemisphere in the temporal fusiform cortex, angular gyrus and lateral occipital cortex. The Intelligibility contrast (Figure 2E) produced similar activation to the Phonology contrast, bilaterally in the middle and superior temporal gyrus (anterior and posterior) and left frontal orbital cortex. Activation extended anteriorly along the left temporal gyrus and into Broca’s area. The Semantics contrast (Figure 2F) also showed bilateral activation in the auditory cortices (middle and superior temporal gyrus including Heschl’s gyrus and planum temporale) with activation extending along the left temporal fusiform and frontal orbital cortices and right parahippocampal gyrus. Coordinates for the maximum intensity of the activated regions are shown in Supplementary Material Table 2.

**Table 2:**
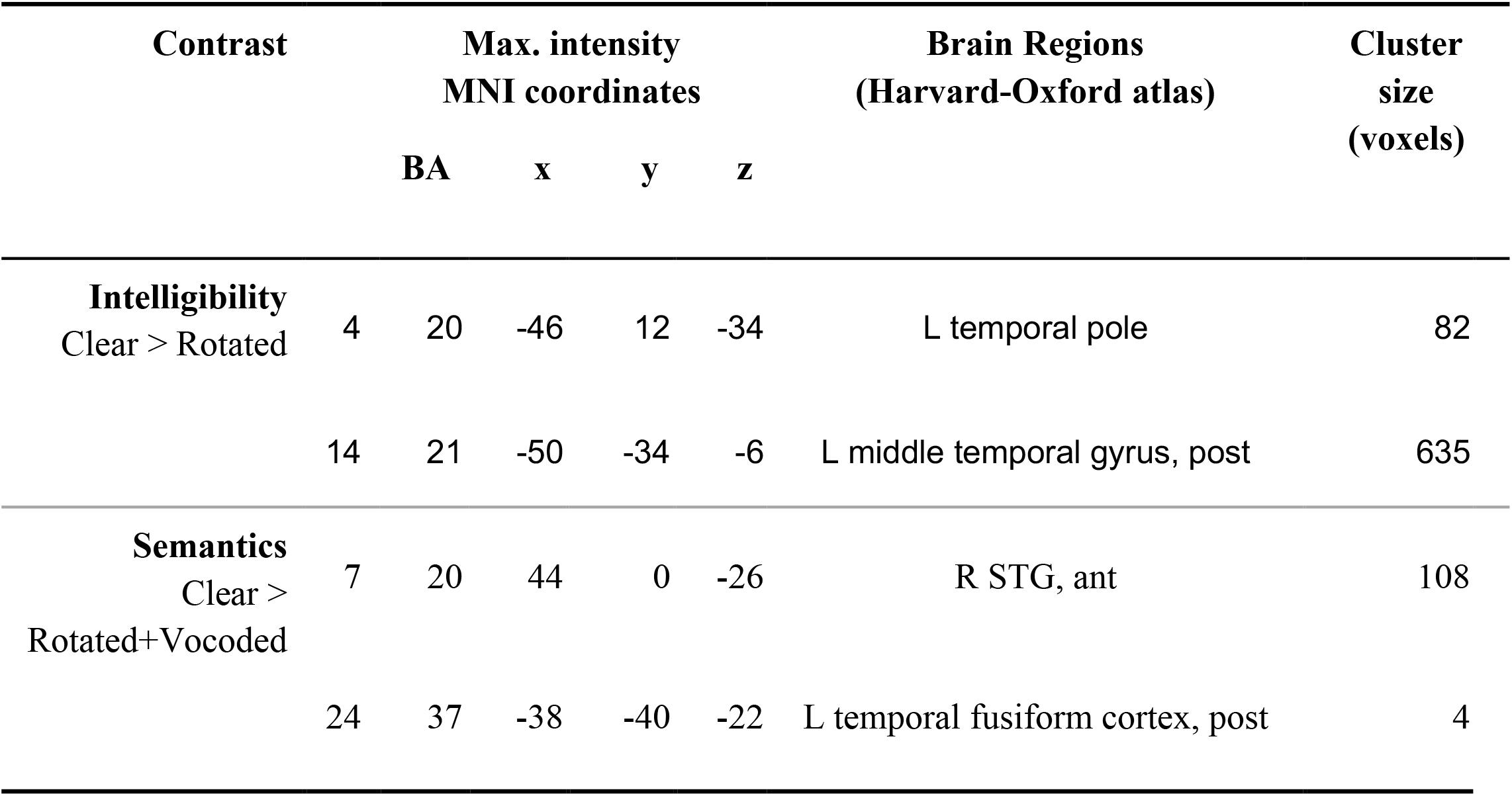
Selected regions of interest (ROIs, from the fMRI task; Fig. 3B and C) used in the rs-fMRI ROI-to-ROI analysis. Clustering threshold = 4.0 voxels. Family-wise error correction = .95. The full table of all ROIs can be found in Supplementary Material Table 3.

**Figure 2:**
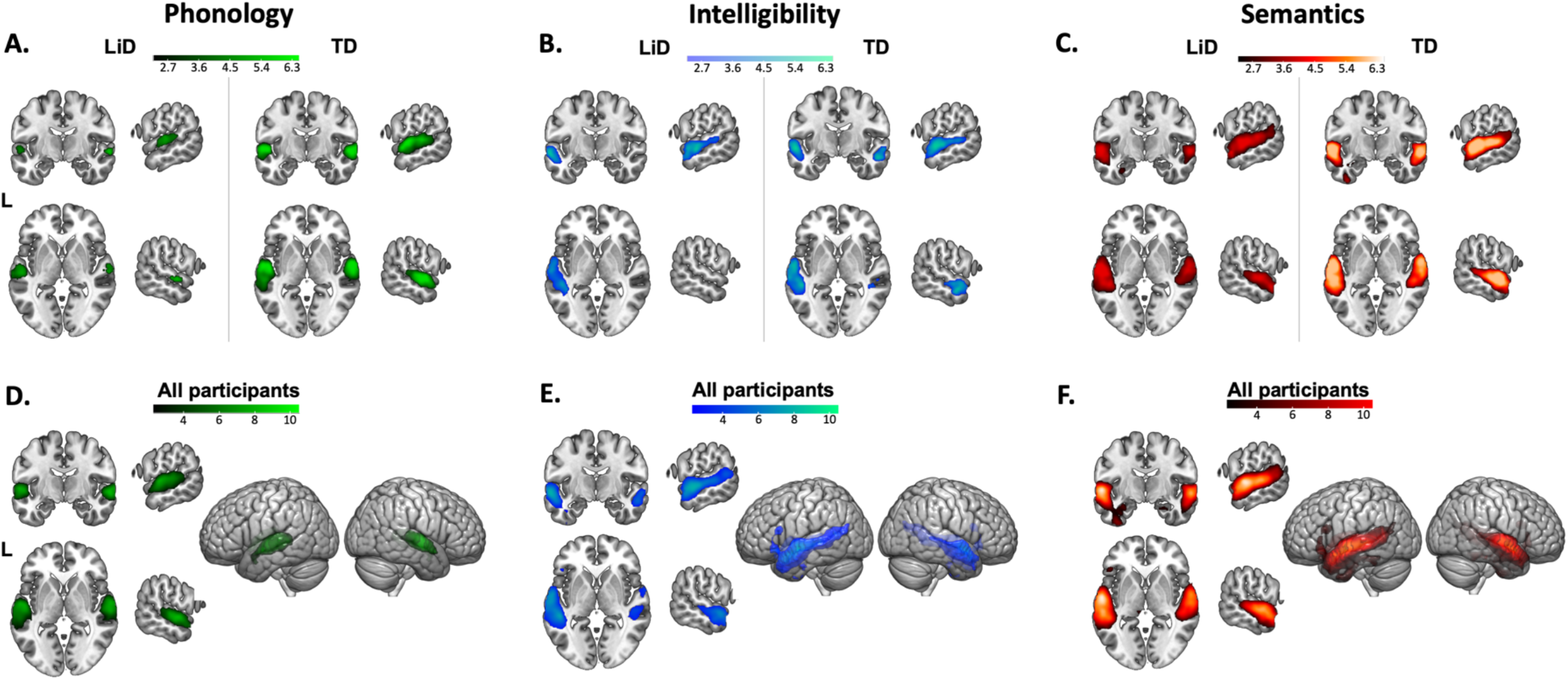
LiDLiD and TD groups showed similar areas of cortical activation in all three contrasts from the fMRI listening task, with no statistical difference between the two groups after correcting for multiple comparisons. Second level GLM analysis for (A-C) groups (clustering threshold = 2.3 voxels, FWE = .95) and (D-F) all participants (clustering threshold = 4.0 voxels, FWE = .95) in the fMRI task, coordinates (±60, −5, 0).Coordinates for maximum intensity voxels for (D-F) can be found in Supplementary Material Table 2. Contrasts are: (A, D) Phonology (green: Rotated > Rotated+Vocoded), (B, E) Intelligibility (blue: Clear > Rotated) and (C, F) Semantics (red: Clear > Rotated+Vocoded). We took the cortical activation across all participants and created parcellated ROIs (Figure 3) for use in the rs-fMRI analysis (figure 4). Images are in neurological orientation. MRIcroGL was used for visualization.

### Cortical functional connectivity

Networks of ROIs suitable for rs-fMRI connectivity analysis were created by parcellating activation produced by the three contrasts from the task-based MRI (Phonology, Intelligibility, and Semantics), combined across groups (Figure 2D-F). The pediatric ADHD-200 sample (Bellec et al., 2017; Craddock et al., 2012) was used as a data-driven, spatially-constrained parcellation method. ROIs smaller than 4 voxels were removed. Cortical activity from the Phonology contrast was divided into a network of 16 ROIs, Intelligibility into a network of 20 ROIs, and Semantics into a network of 24 ROIs (Figure 3A-C; Brodman’s area, maximum intensity coordinates and ROI sizes are in Supplementary Material Table 3). Connectivity was compared between groups (TD = 42, LiD = 39), controlling for age, using a general linear model (GLM) for each of the three networks (Supplementary Material Table 3).

Figure 3 (D-F) shows resting state functional connectivity in the three speech networks, summed across groups. In the Phonology network, each group had connectivity among regions covering bilateral middle and superior temporal gyri, temporal pole, planum temporale, and left planum polare and supramarginal gyrus (Figure 3D). No significant group differences (p-FDR) were found in the Phonology network (Figure 4A, D). Each group had connectivity within the Intelligibility network, covering bilateral middle/superior temporal gyrus and temporal pole along with left pars opercularis, frontal orbital cortex and supramarginal gyrus (Figure 3E). After false discovery rate (FDR) correction, TD children were found to have a significantly stronger temporal correlation between ROIs in left temporal lobe and left middle temporal gyrus (posterior) compared to children with LiD (connection 4-14 in Figure 4B; Table 3). In the Semantics network, both groups of children had connectivity between bilateral middle and superior temporal gyrus, left Heschl’s gyrus, pars triangularis, frontal orbital cortex, planum temporale, temporal fusiform gyrus and right parahippocampal gyrus and planum polare (Figure 3F). Group comparisons showed that, compared to children with LiD, the TD children had stronger temporal correlations between ROIs in the right anterior superior temporal gyrus and the left posterior temporal fusiform cortex (connection 7-24 in Figure 4C; Table 3).

**Table 3:** Network temporal connectivity. Statistical comparison between group differences (TD vs. LiD, corrected for age) for the Intelligibility and Semantics networks (Fig. 4D). No significant group difference was found in the Phonology network. False discovery rate (FDR) was used for multiple comparison correction.

| Contrast | ROI # – ROI #<br>(Fig. 3) | t (78) | p-FDR |
| --- | --- | --- | --- |
| Intelligibility | 4 – 14 | 3.11 | 0.026 |
| Semantics | 7 - 24 | 3.54 | < 0.001 |

**Figure 3:**
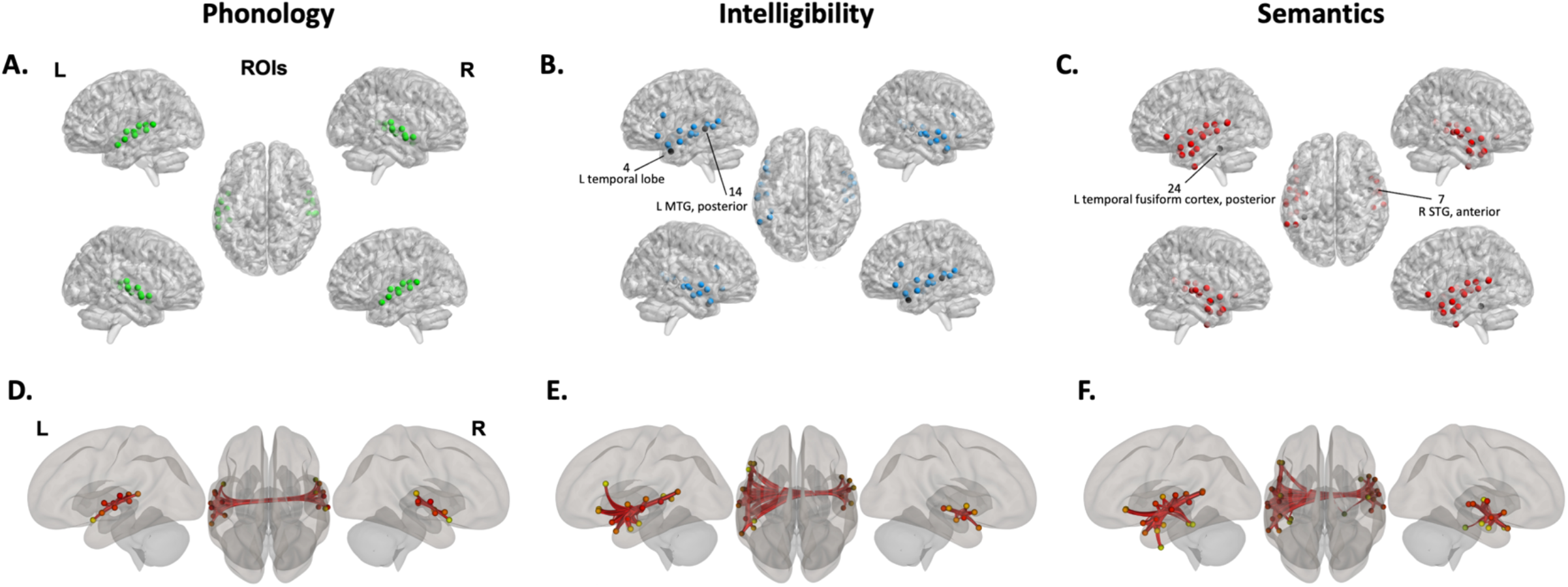
ROIs used for the rs-functional connectivity and their networks across all participants. The cortical activity across all participants in the fMRI task (see Figure 2 D-F) covered large areas and so they were parcellated into smaller ROIs (A-C) for the rs-functional connectivity analysis by applying data-driven spatially constrained parcellation to the areas of activation from the fMRI sentence recognition task using the pediatric ADHD-200 sample. The red lines (D-F) indicate the ROI-to-ROI connections analyzed in each network. Maximum intensity coordinates can be found in Supplementary Material Table 3. Networks are: (A, D) Phonology (green: Rotated > Rotated+Vocoded), (B, E) Intelligibility (blue: Clear > Rotated) and (C, F) Semantics (red: Clear > Rotated+Vocoded). Images are in neurological orientation. For visualization, BrainNet software was used to display foci in (A-C) (Xia, Wang & He, 2013).

**Figure 4:**
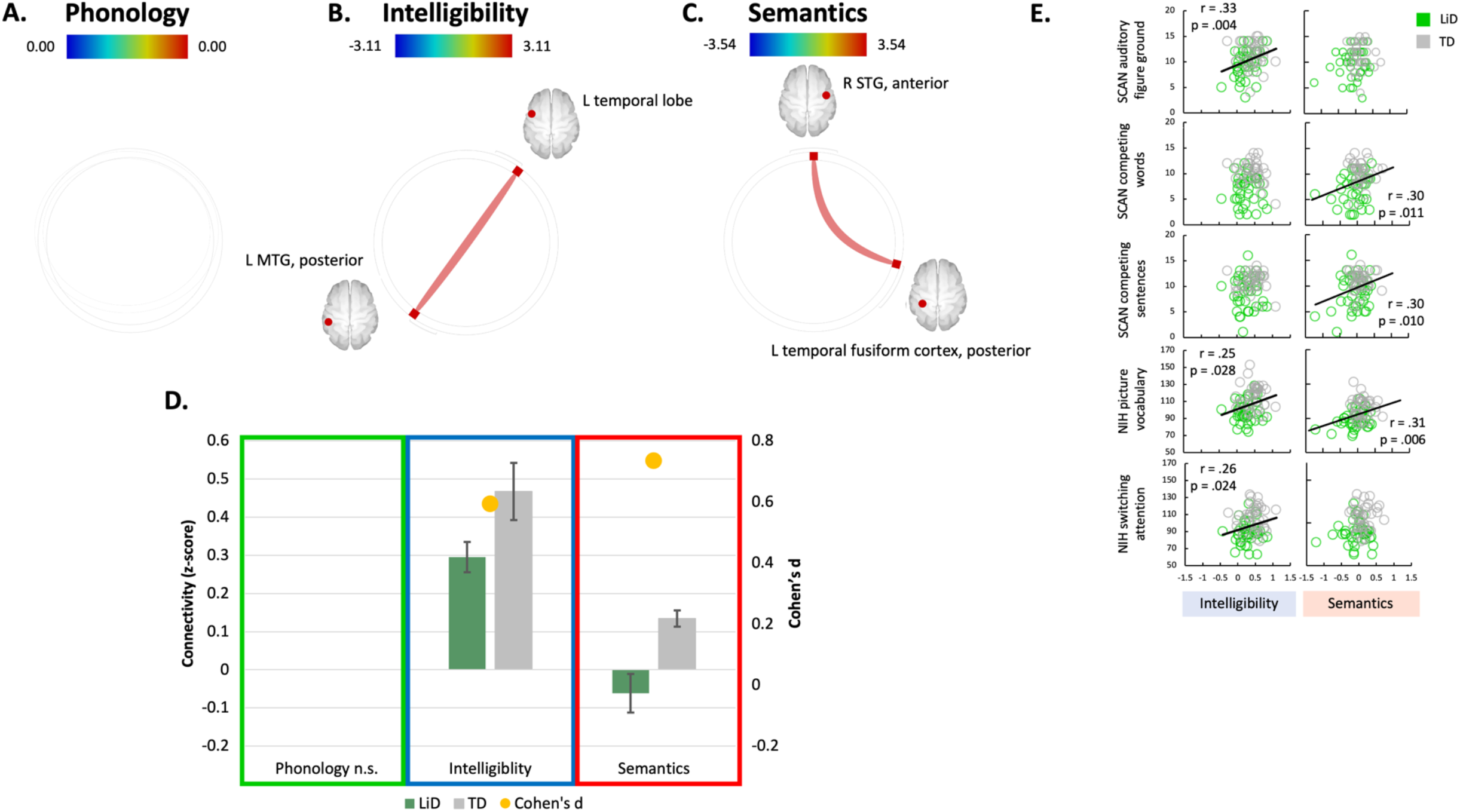
ROI-to-ROI resting state connectivity: the difference between the groups’ listening networks grew from no statistical differences in the phonology network to a minor difference in the intelligibility network to more widespread differences in the semantics network. (A-C) The group comparisons without the effect of age for (A) Phonology, (B) Intelligibility and (C) Semantics networks. Thicker, more saturated color lines represent stronger connections between cortical areas. Note that the colored bar connectivity z score scales vary slightly between connectivity wheels. (D) Details of the ROI-to-ROI connectivity values (left axis) for each group (LiD green, TD grey) and effect sizes (yellow marker, right axis) of the group comparisons without the effect of age. The connections plotted are the ones highlighted as having a significant group difference in the GLM comparing groups without the effect of age (B and C). Connections show the TD group as having significantly stronger connectivity than the LiD group. (E) Scatter plots of brain and behavioral scores demonstrate the correlated patterns of connectomic and behavioural features. Coloured dots in each panel indicate the participant group (LiD: green, TD: grey). The solid black line marks significant correlations (see Supplementary Table 4 for correlation details).

### Relation of cortical functional connectivity to behavioral measures

We explored how the functional connectivity of anatomically separate cortical areas related to behavioral tasks assessing speech in noise ability, dichotic listening and cognition. As shown by Petley et al. (2021), performance on those behavioural tasks was significantly better in the TD than in the LiD group.

The Intelligibility connection had a series of significant positive correlations with speech-in-noise ability (SCAN: auditory figure ground), vocabulary and task switching, a form of executive function (NIH Cognition Toolbox). In contrast, the Semantics connection, involving the left posterior temporal fusiform cortex (7-24; Figure 4C), had significant positive correlations with dichotic listening ability (SCAN: competing words and sentences) and vocabulary (NIH Cognition Toolbox). Interpretation of direction of functional connectivity is ambiguous; an increase in strength of connectivity does not necessarily mean improved ability (Parente et al., 2018). However, these correlations show that, as the children’s behavioural test scores improved, the connection strength within the Intelligibility and Semantic networks increased. The remaining behavioral measures did not relate significantly to connectivity (Figure 4E, Supplementary Material Table 4 and Figure 1).

## DISCUSSION

This study compared TD children and children with LiD on fMRI measures of cortical responses to speech stimuli that assessed processing at the levels of phonology, intelligibility and semantics. The participant groups showed similar cortical activation, with all three sentence contrasts eliciting bilateral activation across the auditory cortex (e.g. Heschl’s gyrus, superior temporal lobe). Activation extended bilaterally into areas thought to be involved in memory and language (e.g., Broca’s area, right parahippocampal gyrus, left temporal fusiform cortex) as the sentence contrasts progressed from emphasizing phonology to intelligibility to semantics. These findings provide support for the hypothesis that primary auditory function does not differ between the two groups when listening to speech. The findings did not support our prediction that children with LiD would show less activation in the speech processing areas than those in the TD group. However, activation was examined using BOLD responses which do not capture the excitatory and inhibitory balance or temporal relationships between cortical areas.

Analysis of the separate rs-fMRI data showed that the TD and LiD groups had different rsMRI responses for the three sentence contrasts. The children with LiD showed less functional connectivity as the sentence contrasts progressed from emphasizing phonology to intelligibility to semantics. This finding supported our functional connectivity hypothesis, but only in networks required for processing speech intelligibility and semantics. It also supports a novel interpretation of childhood LiD as being less of a phonological disorder and more of a language disorder affecting intelligibility and semantics.

Behavioural measures of performance were consistent with the MRI observations in two areas. First was the absence of a significant correlation between functional connectivity for intelligibility and semantic processing in the cortex and the behavioural measure of spatial advantage (the ability to use differences in speaker location to enhance speech intelligibility in noise; LiSN-S), thought to assess auditory processing independently of language and cognition. Second was the significant correlations between functional connectivity for intelligibility and semantic processing with the behavioural measures auditory figure-ground (the ability to hear words-in-noise; SCAN filtered words), dichotic listening (SCAN competing words and SCAN competing sentences), vocabulary (NIH picture vocabulary) and switching attention (NIH switching attention), all thought to assess more cognitively-dominant processes that rely on language and executive functions (Petley et al., 2021). Overall, these data were consistent with the hypothesis that children with LiD have primarily cognitive and language processing deficits rather than auditory processing deficits.

### Bilateralization of speech in 6-12 year old children

Using simple but complete sentence stimuli, we consistently found bilateral activation in children for all speech listening contrasts. After FDR correction, this activation did not significantly differ between groups. These findings suggest that children with LiD use the same cortical areas as TD children for increasingly complex speech processing competencies. However, it is possible that multi-voxel pattern analysis may find finer group differences in the hierarchy for speech processing (e.g., Okada et al., 2010). While the same cortical areas are used by the children with LiD, it is possible that they do so on a different time frame from the TD children. Unfortunately, fMRI does not provide sufficient time resolution to address this possibility.

Our results support the theory of bilateral processing of speech listening in 6-12 year old children. Lateralization of speech processing in adults has been debated across and within imaging modalities (fMRI see Evans & McGettigan, 2017; EEG e.g., Assaneo et al., 2019). For example, Albouy et al. (2020) discussed how acoustic structure leads to a left bias for fast modulation (speech) and a right bias for slow modulation (music) and Rauschecker & Scott’s (2009) unilateral model suggested that the left anterior STG is the hub of successful speech perception. In contrast, Hickok and Poeppel’s (2000) bilateral model proposed a perceptual pathway in each hemisphere processing speech sounds up to the level of semantics (Hickok & Poeppel, 2007). A middle ground has been proposed by Peelle (2012), with right hemisphere dominated activation for “unconnected” speech (i.e., phonemes, syllables, single words) and left hemisphere dominated activation for “connected” speech (i.e., phrases, sentences and narratives).

Bilateral activation reported here differs from the findings of <u>Scott et al. (2000)</u> who reported a left lateralized pathway for speech comprehension using PET, a silent scanning technique. Our results extend those of Halai et al. (2015) who found bilateral activation during continuous fMRI scanning. Both these studies tested young adults and used similar stimuli to those used here. However, speech presentation was continuous, and passive in the sense that the participants did not perform any task during scanning. Rather, comprehension was assessed after scanning and outside the scanning room. In contrast, we required children to provide a behavioural response to each short sentence presentation to encourage attention and as a metric of attention. We additionally used a silent, “sparse” acquisition protocol (Hall et al., 1999; Vannest et al., 2009), in contrast to the previous (Halai et al., 2015) continuous fMRI scanning protocol. Scanner noise superimposed on speech listening has been shown to increase listening effort (Peelle et al., 2010) and to engage additional and or different cortical areas compared with silent acquisition protocols. For example, in a meta-analysis of 57 speech comprehension studies, Adank (2012) showed that continuous scanning more strongly activated regions of the supplementary motor area and anterior cingulate gyrus, while sparse scanning showed more extensive activation in the STS.

### Functional connectivity: Brain and behaviour

We used temporal correlations in rs-fMRI to compare how cortical areas associated with speech processing competencies worked together in children with and without LiD. Phonology is the system of processing the smallest units of speech sounds and their linguistically appropriate combinations. We found no group difference in the connectivity of this network. As we progressed up the speech processing competency hierarchy to the Intelligibility network, we found that temporal correlation between left temporal lobe and left posterior middle temporal gyrus activity was stronger in the TD children. These areas are well known for processing auditory information (temporal lobe) and speech comprehension (left MTG, Acheson & Hagoort, 2013; Dronkers et al., 2004). Posterior MTG has been associated with lexical and semantic access in a sound-to-meaning network (Hickok & Poeppel, 2000, 2004). It is unknown whether stronger or weaker temporal correlations between brain regions is beneficial (Parente et al., 2018). However, relationships found here between the variability of brain connectivity strength and of behaviour provides evidence on this key issue. Children with stronger connectivity performed better on cognitive tasks assessing vocabulary and switching attention. Increased connectivity may thus be indicative of increased processing efficiency and/or suppression of a task-relevant network, in this case language. Note that the behavioural measures used here have been shown to be either little affected by age (audiogram: Hunter et al., 2021) or were standardized across age (ECLiPS, LiSN-S, SCAN and NIH toolbox (Petley et al., 2021).

Group comparisons in the Semantics network highlighted that, compared to children with LiD, the TD children had stronger temporal correlations between auditory areas (right STG) and the left temporal fusiform cortex, associated with word recognition and the recovery of meaning from an impoverished acoustic signal (Davis & Johnsrude, 2003). Brain-behaviour correlations suggest that decreased connectivity between auditory and word recognition cortical areas was associated with impaired speech listening (dichotic listening and picture vocabulary; Table 2). Recently, brain-behaviour correlations have been called into debate due to extremely large datasets being required to reach even very small correlations (Marek et al., 2022). However, we consistently found mild to moderate sized correlations with a peak of ‘r’ = 0.33 (mean ‘r’ = 0.29, first percentile ‘r’ = 0.26; Supplementary Material Table 4).

The cortical areas highlighted in this study (Figure 4 B and C) draw focus to language production, memory encoding and retrieval, and word recognition (Acheson & Hagoort, 2013; Davis & Johnsrude, 2003; Dronkers et al., 2004; Hickok & Poeppel, 2000, 2004) as target areas of further research in children with LiD. Group differences were found in networks of latter stages of speech processing competencies, in the intelligibility and semantics networks. Specifically, between the MTG/STG and the temporal lobe. The reduction in connectivity in these networks suggest that childhood LiD is a semantic and intelligibility disorder. However, no group differences were found in the phonological networks, an early stage of speech processing competencies, suggesting that childhood LiD is not a phonological disorder.

Further assessment where the ‘break’ in speech listening occurs in LiD would provide a clearer avenue for research into effective evidence-based treatments. Future studies could utilize paradigms with anomalous and mispronounced words, with imaging techniques used in parallel to assess if the type of speech processing errors during such tasks are affected by LiD. For example, was it detected when presented (i.e. when judging it’s intelligibility) or did the listener reach the end of the sentence and have to ‘go back’ when they are unable to connect meaning to the sentence. Complementary, time-sensitive techniques (EEG, MEG) could also investigate whether this difficulty is due to a bottleneck in processing leading to increased listening effort, cognitive effort or fatigue (McGarrigle et al., 2014). As LiD may build up during a listening event (i.e., over several minutes; Roebuck & Barry, 2018), it is important to assess the children’s ability throughout the task with cross-sectional time points rather than using summary values of the complete listening event (McGarrigle et al., 2020).

### Relationship to neurodevelopmental disorders and cognitive function

Children identified with LiD have difficulty in speech listening compared to TD children (Petley, Hunter, Motlagh Zadeh, et al., 2021). These groups were further distinguished here by functional connectivity differences between cortical areas associated with language production, memory and word recognition, rather than by the activity or connectivity of primary auditory cortical areas. However, at least 50% of children referred/diagnosed with auditory processing disorder (APD; (British Society of Audiology, 2018; Dillon & Cameron, 2021; Jerger & Musiek, 2000), a clinical label closely related to LiD (Dillon & Cameron, 2021), have also been diagnosed with developmental language disorder (DLD), dyslexia/reading disorders, attention deficit/hyperactivity disorder (ADHD) or more than one of these other neurodevelopmental disorders (Dawes & Bishop, 2010; Ferguson et al., 2011; Gokula et al., 2019; Moore et al., 2018; Sharma et al., 2009). This high level of comorbidity was echoed in the study reported here. While LiD was the primary referral for the study, as with other neurodevelopmental disorders it does not stand in isolation. A background caregiver questionnaire showed that half of the children with LiD also reported a diagnosis of ADHD, 9% autism spectrum disorders and 26% had seen a speech language pathologist. However, these data were not entered into the analysis as we did not conduct gold standard assessments for these other developmental disorders. We are currently using a web-based resource (Neurosynth; Yarkoni et al., 2011) that allows functional connectivity analysis of brain areas defined by a meta-analysis of published fMRI activation coordinates to explore whether there are shared audio-based neurological patterns in children with primary diagnoses of other developmental disorders, such as ASD and ADHD, with LiD.

Further investigation into the neurodevelopmental basis of LiD may particularly inform the understanding of language disorders. Our results show typical phonology but impaired non-phonological speech connectivity in children with LiD. This differs from DLD, which presents with both abilities impaired, and dyslexia, which presents with impaired phonological and typical non-phonological abilities in reading (Bishop & Snowling, 2004; Delage & Durrleman, 2018). It is possible that altered cortical language processing leads to LiD. However, it could be that altered cortical language processing may be a consequence of LiD. Further longitudinal analysis of these children is designed in part to address these issues.

The neuroimaging results presented here highlight the importance of non-auditory factors, specifically language, in audiological testing at a cortical level. There is a growing recognition of the importance of language and, specifically, speech-in-noise (SiN) intelligibility in everyday hearing (Dillon & Cameron, 2021; Golestani et al., 2009; Killion et al., 2004; Magimairaj et al., 2021; Smits et al., 2013). It has been proposed that such testing could supplement, or even replace pure tone detection as an audiometric gold standard (Hewitt, 2018). However, both SiN test instructions and test items pose a challenge to language and memory as well as auditory function. While those cognitive aspects of auditory testing and learning have previously been dismissed as procedural issues (Hawkey et al., 2004), they are an intimate component of a SiN test. These neuroimaging results provide insight into mechanisms of how the advanced stages of speech processing where auditory information is translated into language during speech perception may be disrupted in LiD, a common form of auditory impairment in both children (Moore et al., 2018) and adults (Edwards, 2020). They also add to a growing literature on the role of cognitive function in hearing (Moore et al., 2014; Rönnberg et al., 2013; Sharma et al., 2019; B. Shinn-Cunningham, 2017).

### Conclusions

Our results provide the first multifaceted neurological profile for children classified with LiD, based on caregiver report and normal peripheral auditory function. Children with LiD recruited the same cortical areas as their peers when processing increasing complexities of speech. However, how these cortical areas work together does differ between the two groups. Differences in functional connectivity were found at the more advanced stages of speech listening where the intelligibility and semantics of the speech are processed, specifically in the left temporal lobe, posterior middle temporal gyrus, posterior temporal fusiform cortex and right superior temporal gyrus. These highlighted cortical connections related to the children’s behavioural abilities in dichotic listening, speech-in-noise, attention, memory and verbal vocabulary abilities. Overall, the data provide support for the hypothesis that children with LID are primarily affected by cognitive and, particularly, language processing deficits.

## METHODS

### Participants

Eighty-five participants aged 6-12 years completed the fMRI sentence task and 81 participants completed the rs-fMRI (see Table 1). Seventy-six of participants who completed the fMRI sentence task also completed the rs-fMRI. All participants had normal audiometric hearing with thresholds < 25 dB HL at all octave-interval frequencies from 0.25 - 8 kHz in both ears (Figure 1A). In this paper we focus on the fMRI and its relationship with behavioural responses from the baseline of our longitudinal ‘SICLID’ study examining correlates of LiD in children. Extensive analysis of ear function and behavioural responses of these children is reported elsewhere (Hunter et al., 2021, 2022; Petley et al., 2021).

Caregivers of all participants completed a standardized, validated and reliable checklist of everyday listening and related skills (ECLiPS; Barry & Moore, 2021). Children recruited through advertisement all scored < 10th percentile of ECLiPS standardized scores. Additional children with an audiological diagnosis of auditory processing disorder (APD; n=14) were placed in the LiD group. All but one of these also scored < 10th percentile on the ECLiPS (Figure 1C). Participants scoring within the upper 90^th^ percentile on the ECLiPS and with no history of developmental disorders or delays were classified as TD (further details in Petley et al., 2021).

Eligibility for both groups included English as the child’s native language, and a reported absence of otologic, neurologic or psychiatric disease, or of intellectual insufficiency that would prevent or restrict their ability to complete testing procedures. TD participants were additionally required to have no known history of developmental delay, or attention or language disorder. While half of the children with LiD also reported a diagnosis of ADHD, 9% autism spectrum disorders and 26% had seen a speech language pathologist. Eligibility was determined based on caregiver responses on a medical and educational history ‘Background’ questionnaire (Supplementary Material Table 1).

This study was approved by the Institutional Review Board of Cincinnati Children’s Hospital (CCH) Research Foundation. Prior to completion of study-related imaging and behavioural testing, caregivers reviewed the informed consent form with a study staff member. Children aged 11 and above were also assented using a child-friendly version of the consent document, per institutional policy. All participants received financial compensation for their participation.

### MRI acquisition

MRI was performed via a 3T Philips Ingenia scanner with a 64-channel head coil and Avotec audiovisual system. All participants were awake throughout the scanning. The protocol was modified from a previous large, cross-sectional examination of brain function (Holland & Vannest, 2015) and included a high-resolution T1-weighted anatomical scan, fMRI sentence task (4.9 minutes) and rs-fMRI (5 minutes). The fMRI sentence task was acquired with a sparse scanning protocol (‘HUSH’, details below); TR/TE = 2000/30 ms, voxel size = 2.5 × 2.5 × 3.5 mm, 39 axial slices. A total of 147 volumes was acquired by alternating scanning for 6 seconds (3 volumes) and not scanning for 6 seconds, 49 times. Cardiac and respiration signals were collected during the fMRI sentence task using the scanner’s wireless respirator bellows and Peripheral Pulse Oximeter. The rs-fMRI acquisition was acquired with TR/TE = 2000/30 ms, voxel size = 2.5 × 2.5 × 3.5 mm, 39 axial slices in ascending slice order and 150 volumes. The high-resolution T1-weighted anatomical scan was acquired with TR/TE=8.1/3.7 ms, FOV 25.6 × 25.6 × 16.0 cm, matrix 256 × 256 and slice thickness = 1mm.

### fMRI task

With sound levels reaching 118.4 ± 1.3 dB (A) in a 3T MRI system (Price et al., 2001) special considerations must be made when planning an auditory-based MRI study. In order to protect the participant from the loud environment, foam ear plugs and MRI safe circumaural headphones were worn. The scanner noise may also produce masking of the desired stimuli. Therefore, in the fMRI task we used a ‘Hemodynamics Unrelated to Sounds of Hardware’ (HUSH) scanning protocol (Deshpande et al., 2016; Edmister et al., 1999; Hall et al., 1999; Schmithorst & Holland, 2004) - a sparse temporal sampling protocol where there was no gradient coil noise during presentation of the auditory stimuli. Stimulus levels were elevated to produce adequate signal/noise ratios for accurate responding. We also used a talker identification task instead of a speech recognition task. Instead of asking the children *what* they heard, we asked them *who* had said it (Fig. 1). The children responded with button presses throughout the task so we could ensure they maintained attention to the task.

Sixteen linguistically simple BKB sentences (Bench et al., 1979), designed to be familiar to young children, were recorded by a single male North American speaker, mirroring the paradigm used by Scott et al. (2000) and Halai et al. (2015). These were the Clear speech stimuli (e.g. Figure 1E). Rotated speech stimuli were created by rotating each sentence spectrally around 2 kHz using the Blesser (1972) technique. Rotated speech was not intelligible, though some phonetic features and some of the original intonation was preserved. Rotated+Vocoded speech stimuli were created by applying 6 band noise-vocoding (Shannon et al., 1995) to the Rotated speech stimuli. While the Rotated+Vocoded speech was completely unintelligible, the character of the envelope and some spectral detail was preserved.

Participants were told that they would be completing a matching game where they would hear a sentence and then see a picture (of a “man” or an “alien”). If the picture matched who said the sentence (man - Clear speech, alien - Rotated or Rotated+Vocoded speech), the participant pressed a button with their right thumb, if the picture did not match, they pressed a second button with their left thumb. The participants were asked to respond as fast and as accurately as possible. A sticker was placed on the participants’ right hand to provide a reminder as to which hand was correct for matching voice and picture.

Before scanning, each participant was familiarized with the sentence task and completed three practice trials with verbal feedback from the tester. If a trial was completed incorrectly, the stimuli and instructions were reintroduced until the participant showed understanding. During scanning, each participant completed 48 matching trials, 16 of each sentence type, with no feedback. To maintain scanner timings the behavioural task continued regardless of whether the participant responded. However, if the child did not press a response button on three trials in a row the tester provided reminders/encouragement over the scanner intercom between stimulus presentations.

### fMRI data analysis

First-level fMRI data were processed using FSL (FMRIB Software Library, https://fsl.fmrib.ox.ac.uk/fsl/). The T1 brain data were extracted using BET and normalized and resampled to the 2 mm isotropic MNI ICBM 152 non-linear 6th generation template using FLIRT. For the sparse HUSH acquisition, the volumes were separated and combined into three files according to the volume’s order during the scanner-on period. Each of the three files was pre-processed separately and first-level statistics computed. The three statistical images were then averaged together using a one sample t-test. This was done to account for the difference in intensity among the volumes due to T2* relaxation effects. The pre-processing steps included the following. FSL’s BET was used for brain extraction of the functional data. Outlying functional volumes were detected with ‘fsl_motion_outliers’ using the RMS intensity difference metric. AFNI’s ‘3dretroicor’ was used to regress out the cardiac and respiration signals using a RETROICOR approach (Glover et al., 2000). Motion correction was carried out by MCFLIRT. A GLM was used to regress motion-related artifacts from the data using 6 regressors for the motion parameters and an additional regressor for each outlying volume. The amount of motion during the scans (the number of outlying volumes for each participant) did not differ between groups, *p* = .62 (Table 1). The data were spatially smoothed using a Gaussian kernel with a sigma of 3 mm and temporally filtered with a high pass filter with a sigma of 30 seconds. The results were interpolated to a 2 mm isotropic voxel size and aligned to the Montreal Neurological Institute (MNI) template by first co-registering it with the participant’s T1 using FSL’s FLIRT.

Second-level analysis was also conducted using FSL. A GLM approach was used to create group activation maps based on contrasts between conditions for all participants (i.e. regardless of LiD/TD status) with age as a covariate. Group composite images were thresholded using a family-wise error correction (p < 0.05) and clustering threshold of k = 4 voxels. Three BOLD activation contrasts were used to search for brain loci responding to different aspects of listening to language (Halai et al., 2015; modified from Scott, 2000). First, a ‘Semantics’ activation map, whereby the signal with intelligibility, intonation, phonetics, and prosody was contrasted with one lacking all of these attributes except prosody (Clear > Rotated+Vocoded). Second, an ‘Intelligibility’ activation map contrasted the signal with all speech attributes to one retaining intonation, phonetics and prosody (Clear > Rotated). Third, a ‘Phonetics’ activation map contrasted a signal with intonation, phonetics and prosody with one having only prosody (Rotated > Rotated+Vocoded).

Behavioural responses from the fMRI task were assessed using a 2 (group: TD, LiD) ✕ 3 (sentence type: Clear, Rotated, Rotated+Vocoded) repeated measures analysis of variance (ANOVA) for accuracy and again for RT (Figure 1F). Where the assumption of sphericity was violated, degrees of freedom were corrected using Greenhouse-Geisser estimates of sphericity.

### Resting state fMRI

During this second scan the participant was asked to lie still, keep their eyes open and look at the (central) white cross on the black screen. During the 5 minute period they were not performing an exogenous task. Eyes were monitored by the tester through CCTV and no child fell asleep during the task.

For the rs-fMRI scan, pre-processing and analysis was performed in the CONN toolbox using standard spatial and temporal pipelines (Whitfield-Gabrieli & Nieto-Castanon, 2012). For spatial smoothing a FWHM of 8mm was used. The Artifact Detection Tool (ART, https://www.nitrc.org/projects/artifact_detect) within CONN was used to regress out framewise motion. The number of frames regressed out was compared between groups with no significant group differences, *p* = .44 (Table 1).

### Resting state ROI-to-ROI analysis

The group activation maps from the fMRI task’s three contrasts (cluster threshold = 4 voxels; p-FWE = .95) were used to define the ROIs of advancing speech processing competency networks (Phonology, Intelligibility and Semantics). However, as these areas of activation were large (e.g. an area of 4351 voxels in the left frontal lobe as part of the Semantics network, Supplementary Material Table 2), we applied the parcellation from the pediatric ADHD-200 sample (Bellec et al., 2017) to each network. This created smaller and more appropriate ROIs for connectivity analysis of each network (Figure 3D-F, Supplementary Material Table 3). ROIs smaller than 4 voxels were not included in the analysis.

Conn was used to test the functional relationship between each pair of ROIs identified in the fMRI sentence listening task. The mean time course of all voxels within each ROI was used to calculate individual pairwise Pearson correlations. The *r* values were normalized to *z* values via Fisher’s z-transformation. We then used these *z* values to explore the relationship between the three listening networks and behavioural measures. Statistical thresholds were set to p < .05 (corrected) at the single voxel level, and the resulting connections were thresholded at seed-level by intensity with FDR correction (p < .05).

### Caregiver questionnaire

#### *Everyday listening skills – ECLiPS* (Barry & Moore, 2021)

The ECLiPS is a standardized caregiver-report measure of listening and communication difficulties. Caregivers rated 38 simple statements about their child on a five-point scale, ranging from strongly disagree to strongly agree.

### Behavioral measures

Resting state temporal connections with significant group differences were correlated with behavioural measures described briefly below (see Petley et al., 2021 for further detail and data). Study data were collected and managed using REDCap electronic data capture tools hosted at Cincinnati Children’s Hospital (Harris et al., 2009, 2019). REDCap (Research Electronic Data Capture) is a secure, web-based software platform designed to support data capture for research studies, providing 1) an intuitive interface for validated data capture; 2) audit trails for tracking data manipulation and export procedures; 3) automated export procedures for seamless data downloads to common statistical packages; and 4) procedures for data integration and interoperability with external sources.

### Listening in Spatialized Noise-Sentences

(Brown et al., 2010; Cameron & Dillon, 2007, 2009) LiSN-S (US version) is a standardized test assessing speech in noise ability. Binaural target (T) sentences were presented through headphones along with two other distracting sentences (D1, D2). The children were asked to repeat the sentences of the target voice only. Distracting sentences remained constant at 55 db SPL. After each correct trial the target voice descended in level (4 dB), but if the child incorrectly repeated back over 50% of the sentence the level increased (by 2 dB).

Four listening conditions are made by manipulating D1 and D2 with respect to T (same voice, different voices; same direction, 0°, different direction, ± 90° azimuth). Three difference (“Advantage”) scores are calculated to reduce the influence of language and cognitive demands of each condition (Dillon et al., 2014): Talker Advantage (different voices - same voice); Spatial Advantage (different directions – same direction); and Total Advantage (different voices and directions – same voices and directions). LISN-S software calculated these difference scores for each participant.

#### *SCAN-3:C* (Keith, 2009; 2000)

SCAN-3:C is a US-standardized test battery often used by audiologists to diagnose APD in children (Emanuel et al., 2011). We used the task to obtain comparison measures relative to other studies of auditory processing ability (e.g. Kelley & Littenberg, 2019), but not to group children as LiD or TD. Subtests used in our battery were Auditory Figure Ground - assessing the ability to repeat words presented against background multi-talker speech; Competing words - a dichotic listening task where the child repeats different words presented simultaneously to each ear, repeating that from a designated ear first; Filtered words - assessing ability to identify words that are low pass filtered at 750 Hz; and Competing Sentences - a dichotic listening task where different sentences are presented simultaneously to each ear, and the child is asked to repeat the sentence from a designated ear. Both subtest and a standardized SCAN composite score are calculated.

#### Cognition - NIH toolbox (Weintraub et al., 2013)

The NIH toolbox - Cognition Battery is a collection of US-standardized tests from which we used measures of selective attention (Flanker Inhibitory Control and Attention Test), episodic memory (Picture Sequence Memory Test), executive function (Dimensional Change Card Sort Test) and picture vocabulary. Each visually administered test took 5 - 15 minutes to complete on an iPad. Age-corrected subtest and an overall ‘early childhood composite’ scores were calculated for each participant.

## Data Availability

The dataset generated during and analysed during the current study are available from the corresponding author. ROIs used in the rs-fMRI analysis are available at GitHub.

https://github.com/stewarthannahj/ROIs_SICLiD.git

## Abbreviations

CANS: central auditory nervous system
DLD: developmental language disorder
rs-fMRI: resting state-fMRI
FOV: field-of-view
FEW: family wise error correction
GLM: general linear model
HUSH: Hemodynamics Unrelated to Sounds of Hardware
LiD: listening difficulties
MEG: magnetoencephalography
MNI: Montreal Neurological Institute
MTG: middle temporal gyrus
FDR: false detection rate
ROI: region-of-interest
SLI: specific language impairment
STG: superior temporal gyrus
STS: superior temporal sulcus
TD: typical developing
TE: echo time
TMS: transcranial magnetic stimulation
TR: repetition time

## DATA AVAILABILITY

In accordance with ethics requirements, the dataset generated and analysed during the current study are available from the corresponding author. ROIs used in the rs-fMRI analysis are available at GitHub (https://github.com/stewarthannahj/ROIs_SICLiD.git).

## ACKNOWLEDGEMENTS

Thanks to Dr Kim Leikin and Prof. Scott K Holland for aspects of study design and to Prof. Stuart Rosen and Dr Peter Chiu for the scripts to rotate and vocode the stimuli for the fMRI sentence task. Thanks also to Audrey Perdew, Nicholette Sloat and Dr Ronan McGarrigle for assisting in MRI acquisition. Many thanks to the CCH MRI techs (Matt Lanier, Lacey Haas, Kaley Bridgewater, Kelsey Murphy, Byrnne Williams and Marty Jones) for many hours of scanning.

## FUNDING

This research was supported by NIH R01DC014078, NIH 2UL1TR001425 and by Cincinnati Children’s Research Foundation. DRM is supported in part by the NIHR Manchester Biomedical Research Centre.

## COMPETING INTERESTS

The authors report no competing interests.

## CONTRIBUTIONS

HJS - study design, MRI acquisition, MRI preprocessing and analysis, behavioural analysis, figures and tables, manuscript preparation and revisions

EC - behavioural and rs-fMRI analysis, figures, assisted with manuscript preparation and revisions

LLH - study design, manuscript preparation and revisions

TM - fMRI preprocessing, assisted with manuscript preparation

JV - study design, analysis and presentation, manuscript preparation and revisions

DRM - study design and presentation, funding, manuscript preparation and revisions

## Notes

### Competing Interest Statement

The authors have declared no competing interest.

### Author Declarations

Cincinnati Children's Hospital Medical Center Institutional Review Board. Ethical approval given.

### Summary of Updates

Restructure of the introduction and general tidy up.

