## Supplementary materials for "Speech cortical activation and connectivity in typically developing children and those with listening difficulties"

**Abbreviated title**: Cortical speech networks and listening

Hannah J. Stewart [1, 2, 3], Erin K. Cash [2], Lisa L. Hunter [2], Thomas Maloney [4], Jennifer Vannest [2,5] and David R. Moore [2, 6, 7]

1. Division of Psychology and Language Sciences, University College London, London, UK
2. Communication Sciences Research Center, Cincinnati Children's Hospital Medical Center, Cincinnati, Ohio, USA
3. Department of Psychology, Lancaster University, Lancaster, UK
4. Pediatric Neuroimaging Research Consortium, Cincinnati Children's Hospital Medical Center, Cincinnati, Ohio, USA
5. Department of Communication Sciences and Disorders, University of Cincinnati, Ohio, USA
6. Department of Otolaryngology, College of Medicine, University of Cincinnati, Cincinnati, Ohio, USA
7. Manchester Centre for Audiology and Deafness, University of Manchester, Manchester, M13 9PL, UK

**Correspondence**: Hannah J. Stewart
Department of Psychology
Lancaster University
Lancaster UK


**Keywords**: listening difficulties, pediatric, fMRI, resting state, connectivity, speech perception

**Table 1**: Developmental disorders in each group, as reported by the children's caregivers. (ASD: autism spectrum disorder; ADHD: attention deficit hyperactivity disorder; SLP: speech language pathologist).

| Group | N | ASD | ADHD | Dyslexia | Seen a SLP |
|---|---|---|---|---|---|
| LiD | 43 | 4 | 21 | 1 | 11 |
| TD | 41 | 0 | 0 | 0 | 0 |

**Table 2**: Brodmann area (BA) and MNI coordinates for maximum intensity voxels from the fMRI speech listening task contrasts. Cortical activation is across all 85 participants as groups were not significantly different after correcting for multiple comparisons. All three contrasts showed large areas of activation covering the auditory cortex. With activation extending out towards language (Wernicke's), memory (parahippocampal) and decision making (frontal orbital cortex) areas as the contrasts progressed from simpler stages of speech listening (phonology) to more advanced (semantics). Cluster threshold = 4.0 voxels, p-FDR = .95. See Figure 2.

| **Contrast** | **Max. intensity MNI coordinates** | | | | **Brain Regions (Harvard-Oxford atlas)** | **Cluster size (voxels)** |
|---|---|---|---|---|---|---|
| | **BA** | **x** | **y** | **z** | | |
| **Phonology** Rotated > Rotated+Vocoded | 21 | -48 | 0 | -24 | L middle temporal gyrus | 1603 |
| | 21 | 60 | 0 | -16 | R middle temporal gyrus, ant | 1238 |
| | 41 | -40 | -34 | 6 | L planum temporale | 7 |
| **Intelligibility** Clear > Rotated | 20 | -46 | 12 | -34 | L temporal pole | 2567 |
| | 21 | 50 | 8 | -30 | R temporal pole | 1114 |
| | 48 | -50 | 18 | 18 | L inferior frontal gyrus | 6 |
| **Semantics** Clear > Rotated+Vocoded | 20 | -44 | 6 | -34 | L frontal pole | 4351 |
| | 20 | 46 | 8 | -32 | R temporal pole | 2810 |
| | 20 | -36 | -10 | -46 | L temporal fusiform cortex, post | 33 |
| | 38 | -52 | 26 | -10 | L frontal orbital cortex | 25 |
| | 37 | -38 | -40 | -22 | L temporal fusiform cortex, post | 7 |

**Table 3**: Brodmann area (BA) and MNI coordinates for the regions of interest from the fMRI task used in the rs-fMRI ROI-to-ROI analysis. Cluster threshold = 4.0 voxels, p-FDR = .95.

| Contrast | ROI | Max. intensity MNI coordinates BA | x | y | z | Brain Regions (Harvard-Oxford atlas) | Cluster size (voxels) |
|---|---|---|---|---|---|---|---|
| **Phonology** Rotated > Rotated+Vocoded | 1 | 21 | 66 | -18 | -10 | R middle temporal gyrus | 317 |
| | 2 | 22 | -60 | -42 | 6 | L supramarginal gyrus, post | 23 |
| | 3 | 21 | 66 | -4 | -6 | R superior temporal gyrus | 351 |
| | 4 | 21 | -60 | -16 | -10 | L middle temporal gyrus, post | 192 |
| | 5 | 22 | 60 | -28 | 10 | R planum temporale | 5 |
| | 6 | 21 | 52 | -4 | -10 | R superior temporal gyrus, ant | 17 |
| | 7 | 21 | -48 | -2 | -14 | L planum polare | 319 |
| | 8 | 21 | 52 | 4 | -14 | R superior temporal gyrus, ant | 60 |
| | 9 | 21 | -54 | 0 | -10 | L superior temporal gyrus, ant | 153 |
| | 10 | 21 | -48 | 4 | -22 | L temporal pole | 25 |

| | | | | | | | |
|---|---|---|---|---|---|---|---|
| | 11 | 41 | -40 | -34 | 6 | L planum temporale | 19 |
| | 12 | 21 | -60 | -28 | -6 | L middle temporal gyrus, post | 330 |
| | 13 | 21 | 60 | 0 | -16 | R middle temporal gyrus, ant | 356 |
| | 14 | 21 | -68 | -28 | 2 | L superior temporal gyrus, post | 400 |
| | 15 | 21 | -48 | 0 | -24 | L middle temporal gyrus, ant | 149 |
| | 16 | 21 | 60 | -22 | -6 | R middle temporal gyrus, post | 131 |
| **Intelligibility** Clear > Rotated | 1 | 21 | 60 | -20 | -6 | R middle temporal gyrus, post | 11 |
| | 2 | 21 | -60 | -46 | 2 | L middle temporal gyrus, temporoccipital part | 131 |
| | 3 | 21 | 50 | 8 | -30 | R temporal pole | 210 |
| | 4 | 20 | -46 | 12 | -34 | L temporal pole | 82 |
| | 5 | 21 | 66 | -4 | -6 | R superior temporal gyrus, ant | 37 |
| | 6 | 20 | -52 | -18 | -14 | L middle temporal gyrus, post | 208 |
| | 7 | 20 | 44 | 6 | -26 | R temporal pole | 25 |
| | 8 | 21 | -46 | -4 | -22 | L superior temporal gyrus, ant | 255 |
| | 9 | 38 | 58 | 12 | -18 | R temporal pole | 64 |

| | | | | | | | |
|---|---|---|---|---|---|---|---|
| | 10 | 21 | -52 | 0 | -10 | L superior temporal gyrus, ant | 264 |
| | 11 | 20 | -42 | 12 | -28 | L temporal pole | 223 |
| | 12 | 21 | -44 | -40 | 6 | L supramarginal gryus, post | 9 |
| | 13 | 21 | -44 | -48 | -2 | L inferior temporal gyrus | 28 |
| | 14 | 21 | -50 | -34 | -6 | L middle temporal gyrus, post | 635 |
| | 15 | 21 | 52 | 0 | -26 | R middle temporal gyrus, ant | 576 |
| | 16 | 21 | -68 | -28 | 2 | L superior temporal gyrus, post | 116 |
| | 17 | 20 | -48 | -4 | -26 | L middle temporal gyrus, ant | 573 |
| | 18 | 38 | -50 | 24 | -12 | L frontal orbital cortex | 24 |
| | 19 | 20 | 50 | -20 | -16 | R middle temporal gyrus, post | 180 |
| | 20 | 48 | -50 | 18 | 18 | L inferior frontal gyrus, pars opercularis (Broca's) | 6 |
| **Semantics** Clear > Rotated+Vocoded | 1 | 21 | 68 | -20 | -10 | R MTG, post | 301 |
| | 2 | 21 | -52 | -50 | 2 | L middle temporal gyrus, temporoccipital part | 363 |
| | 3 | 20 | 46 | 8 | -32 | R temporal pole | 344 |
| | 4 | 20 | -44 | 6 | -34 | L temporal pole | 185 |
| | 5 | 21 | 66 | -4 | -6 | R superior temporal gyrus, ant. | 420 |

| | | | | | | |
|---|---|---|---|---|---|---|
| 6 | 20 | -52 | -18 | -16 | L middle temporal gyrus, post | 309 |
| 7 | 20 | 44 | 0 | -26 | R STG, ant | 108 |
| 8 | 20 | -46 | -8 | -22 | L MTG, post | 593 |
| 9 | 21 | 48 | 8 | -18 | R temporal pole | 210 |
| 10 | 21 | -52 | 0 | -10 | L superior temporal gyrus, ant | 368 |
| 11 | 20 | -42 | 8 | -30 | L temporal pole | 314 |
| 12 | 21 | -42 | -32 | 2 | L planum temporale | 86 |
| 13 | 21 | -50 | -48 | 2 | L middle temporal gyrus, temporoccipital part | 128 |
| 14 | 21 | -50 | -34 | -8 | L MTG, post | 899 |
| 15 | 21 | 54 | -6 | -26 | R middle temporal gyrus, ant | 792 |
| 16 | 21 | -68 | -28 | 2 | L superior temporal gyrus, post | 419 |
| 17 | 20 | -48 | -6 | -26 | L middle temporal gyrus, ant | 634 |
| 18 | 38 | -52 | 26 | -10 | L inferior frontal gyrus, pars traingularis | 24 |
| 19 | 20 | -48 | -2 | -30 | L middle temporal gyrus, ant | 12 |

| | | | | | | |
|---|---|---|---|---|---|---|
| 20 | 20 | 52 | -18 | -16 | R middle temporal gyrus, post | 625 |
| 21 | 20 | 52 | -6 | -26 | R middle temporal gyrus, ant | 6 |
| 22 | 20 | -44 | -6 | -26 | L inferior frontal gyrus, ant | 10 |
| 23 | 20 | -36 | -10 | -46 | L temporal fusiform cortex | 33 |
| 24 | 37 | -38 | -40 | -22 | L temporal fusiform cortex, post | 4 |

**Table 4**: Mild to moderate correlations were found between temporal connectivity and behavioural outcome measures (auditory processing skills, verbal vocabulary, selective attention, switching attention and episodic memory). Note that only connections with significant group differences (Figure 4) are shown here.

| **Network** | **ROI # – ROI #** | **Behavioral outcome** | **r (77)** | **p** |
|---|---|---|---|---|
| Intelligibility | 14-14 | SCAN auditory figure ground | .33 | .004 |
| | | NIH verbal vocab | .25 | .028 |
| | | NIH task switching | .26 | .024 |
| Speech | 24-7 | SCAN complete words | .30 | .011 |
| | | SCAN complete sentences | .30 | .010 |
| | | NIH verbal vocab | .31 | .006 |
